# Quantitative Prognostic Modeling in Aneurysmal Subarachnoid Hemorrhage: Multicenter Validation of the eSAH Score

**DOI:** 10.64898/2026.07.18.26358390

**Authors:** Saif Salman, Constantin Graf von Moy, Flora Haidenberger, Mahlika Ahmed, Fabian Foettinger, Rohan Sharma, Salvador F Gutierrez-Aguirre, Otavio Frederico de Toledo, Vishal Patel, David Yujia-Wei, Behnam Rezai Jahromi, Nicholas Brandmeir, Kausik Lakkaraju, Mutaz Ombada, Pedro Aguilar-Salinas, David Miller, Bradley Erickson, Ricardo A Hanel, Rabih G Tawk, Richard Byrne, William David Freeman

**Affiliations:** Departments of Neurologic Surgery, Neurology, and Critical Care, Mayo Clinic, Jacksonville, Florida, USA; Department of Neurology, Medical University of Vienna, Vienna, Austria; Comprehensive Center for Clinical Neurosciences and Mental Health, Medical University of Vienna, Vienna, Austria; Neurology, Creighton University, Omaha, Nebraska, USA; Lyerly Neurosurgery, Baptist Neurological Institute, Jacksonville, Florida, USA; Department of Radiology, Mayo Clinic, Jacksonville, Florida, USA; Department of Radiology, Mayo Clinic School of Graduate Medical Education, Mayo Clinic College of Medicine and Science, Rochester, Minnesota, USA; Department of Neurosurgery, University of Helsinki, Helsinki, Finland; Department of Neuroscience, Rockefeller Neuroscience Institute, West Virginia University School of Medicine, Morgantown, West Virginia; Charleston Area Medical Center Institute for Academic Medicine/WVU School of Medicine Charleston campus, Charleston, South Carolina, USA; Department of Radiology, Mayo Clinic, Rochester, Minnesota, USA; Endowed Chair of Stroke and Cerebrovascular Surgery, Baptist Health, Jacksonville, Florida, USA; Chair, Department of Neurologic Surgery, Mayo Clinic, Jacksonville, Florida, USA; Chair, Specialty Neurosurgery Council, Mayo Clinic, Jacksonville, Florida, USA

## Abstract

**Background:** aneurysmal subarachnoid hemorrhage (aSAH) is neurological emergency associated with substantial mortality and disability. Current grading systems such as the modified Fisher Scale (mFS) and World Federation of Neurological Societies (WFNS) score, rely on semiquantitative and examination based assessments. Hence, they demonstrate limited predictive precision. The enhanced subarachnoid hemorrhage (eSAH) score is a simplified quantitative model integrating age, Glasgow Coma Scale (GCS), and cisternal subarachnoid hemorrhage volume (SAHV) to predict clinical outcomes after aSAH.

**Methods:** We performed a retrospective multicenter cohort study that included 1088 patients across three tertiary-care centers the United States. Predictive performance for unfavorable functional outcome, in-hospital mortality and delayed cerebral ischemia (DCI) was evaluated using receiver operating characteristic (ROC) analysis and area under the curve (AUC). Comparative analyses were performed and compared to the WFNS and mFS grading systems.

**Results:** the eSAH score demonstrated excellent discrimination for unfavorable functional outcome at discharge ( AUC 0.89 ) and in-hospital mortality (AUC 0.87). The DCI subscore demonstrated good discriminatory performance for predicting DCI (AUC 0.77). Compared with conventional grading systems, this was superior to both the WFNS (AUC 0.75) and the mFS ( AUC 0.70). increasing eSAH scores were additionally associated with progressively higher rates of mortality and unfavorable functional outcomes.

**Conclusion:** the eSAH score demonstrates strong external validity, reproducibility and superior predictive performance compared with conventional grading systems in a large multicenter cohort. These findings support the clinical utility of quantitative hemorrhage burden integration for early risk stratification in patients with aSAH.

## Introduction

Aneurysmal subarachnoid hemorrhage ( aSAH) accounts for a small proportion of all strokes. However, it contributes disproportionately to neurological mortality and long-term disability^1–3^. Management guidelines emphasize early aneurysm securing, aggressive monitoring and prevention of delayed cerebral ischemia (DCI)^1,4^. Despite advances in healthcare, early risk stratification remains essential to direct clinical decision making and avoid prognostic bias^5^.

Current clinical grading systems such as Hunt and Hess and the World Federation of Neurosurgical Societies (WFNS) scale rely primarily on neurological examination^6,7^. However, such scales can be influenced by various clinical confounders such as sedation, hydrocephalus, metabolic abnormalities, or systemic illness^8,9^. Even patients with favorable WFNS grade may experience poor outcomes^10^. Radiographic grading systems, including the Fisher scale and the modified Fisher Scale, provide a semiquantitative clot burden assessment but lack volumetric precision and demonstrate interobserver variability^6,12^.

Quantitative hemorrhage burden has been consistently associated with outcome and vasospasm risk^12,13^. Automated and semiautomated volumetric methods have demonstrated improved predictive accuracy compared with ordinal scales^7,14^. The recently developed eSAH score integrates age, admission GCS, and the ellipsoid-derived (ABC/2) SAH volume into a simplified 0–5-point model^15^. In the original derivation study, the eSAH score demonstrated strong predictive performance, modified Rankin Scale (mRS) and DCI^15^.

Given that predictive models frequently demonstrate attenuation when applied outside derivation settings, external validation is critical. We therefore evaluated the performance of the eSAH score in an independent tertiary-care US cohort.

## Methods

This retrospective cohort study included 1088 patients with confirmed aSAH treated between 2006 and 2025 across three tertiary centers: Mayo Clinic Rochester (Minnesota, USA), Mayo Clinic Jacksonville (Florida, USA), and the Baptist Neurological Institute Jacksonville (Florida, USA). Patients from the original cohort were excluded to ensure independence^15^.

Institutional review board approval was obtained with a waiver of informed consent. Inclusion criteria required age ≥18 years, confirmed aneurysmal SAH, baseline noncontrast CT scans obtained within 24 hours, and available discharge modified Rankin Scale (mRS). Traumatic SAH and non-aneurysmal hemorrhage were excluded. Patients with missing variables required for eSAH score calculation were also excluded from the analysis.

Cisternal SAH volume was calculated using the ABC/2-derived ellipsoid method^15,16^. The original eSAH scoring threshold^15^ was applied without modification, total score ranged from 0 to 5, incorporating GCS category, SAH volume category, and age. The DCI subscore included only GCS and SAH volume categories.

Primary outcomes were unfavorable discharge and functional outcomes (mRS>3) and in-hospital mortality. The Secondary outcome was DCI, defined according to the established criteria^15^.

Statistical analyses were performed using Python (version 3.14) with the pandas, numpy, scipy, and scikit-learn libraries. Continuous variables are presented as mean (standard deviation) or median (interquartile range) depending on distribution, and categorical variables are reported as counts and percentages.

Comparisons between patients with favorable (mRS 0–3) and unfavorable (mRS 4–6) outcomes were performed using Student’s t-test or the Mann–Whitney U test for continuous variables and the chi-square test for categorical variables.

The predictive performance of the eSAH score for unfavorable outcome and mortality was evaluated using receiver operating characteristic (ROC) curve analysis. Discriminatory ability was quantified using the area under the ROC curve (AUC).

Comparative analyses with other grading scales such as the modified Fisher Scale (mFS) and the World Federation of Neurosurgical Societies (WFNS) score, were included in this cohort and compared to the eSAH score.

A two-sided p-value <0.05 was considered statistically significant.

## Results

A total of 1088 patients met the inclusion criteria. Favorable functional outcome at discharge (mRS 0-3) was observed in 668 patients (61.4%), whereas 420 patients (38.6%) had an unfavorable outcome (mRS 4-6). In-hospital mortality occurred in 186 patients (17.1%).

Baseline clinical characteristics stratified by functional outcome are summarized in Table 1. Patients with unfavorable outcomes were significantly older than those with favorable outcomes (64 vs 56 years, p<0.001). They also presented with markedly lower admission GCS scores (median 7 vs 15, p<0.001) and higher cisternal SAH volumes (12.3 vs 4.5 mL, p<0.001). Delayed cerebral ischemia occurred more frequently in patients with unfavorable outcomes (241 vs 75 patients, p<0.001).

**Table 1:** Baseline Characteristics.

| **Variable** | **Favorable**<br>**(mRS, 0-3)** | **Unfavorable**<br>**(mRS, 4-6)** | **P value** |
| --- | --- | --- | --- |
| Age, mean (SD) | 56.52 (12.70) | 63.03 (12.50) | <0.001 |
| GCS score at admission, median (IQR) | 15 (14-15) | 7 (4-11) | <0.001 |
| SAHV (mL), mean (SD) | 4.69 (2.25) | 13.76 (6.63) | <0.001 |
| DCI, No. | 75 | 241 | <0.001 |

The discriminative performance of the eSAH score was evaluated using ROC curve analysis (Figure 1). The eSAH score demonstrated excellent discrimination for predicting unfavorable functional outcome at discharge with an AUC of 0.89. Similarly, the eSAH score showed excellent performance for predicting in-hospital mortality with an AUC of 0.87. The DCI subscore demonstrated good discriminatory performance for predicting delayed cerebral ischemia with an AUC of 0.77. The eSAH score was also compared with the WFNS and the mFS, and demonstrated superior predictive performance relative to both. The mFS predicted an unfavorable outcome with an AUC of 0.70 while theWFNS score demonstrated an AUC of 0.75 compared with an AUC of 0.89 for the eSAH score.

**Figure 1:**
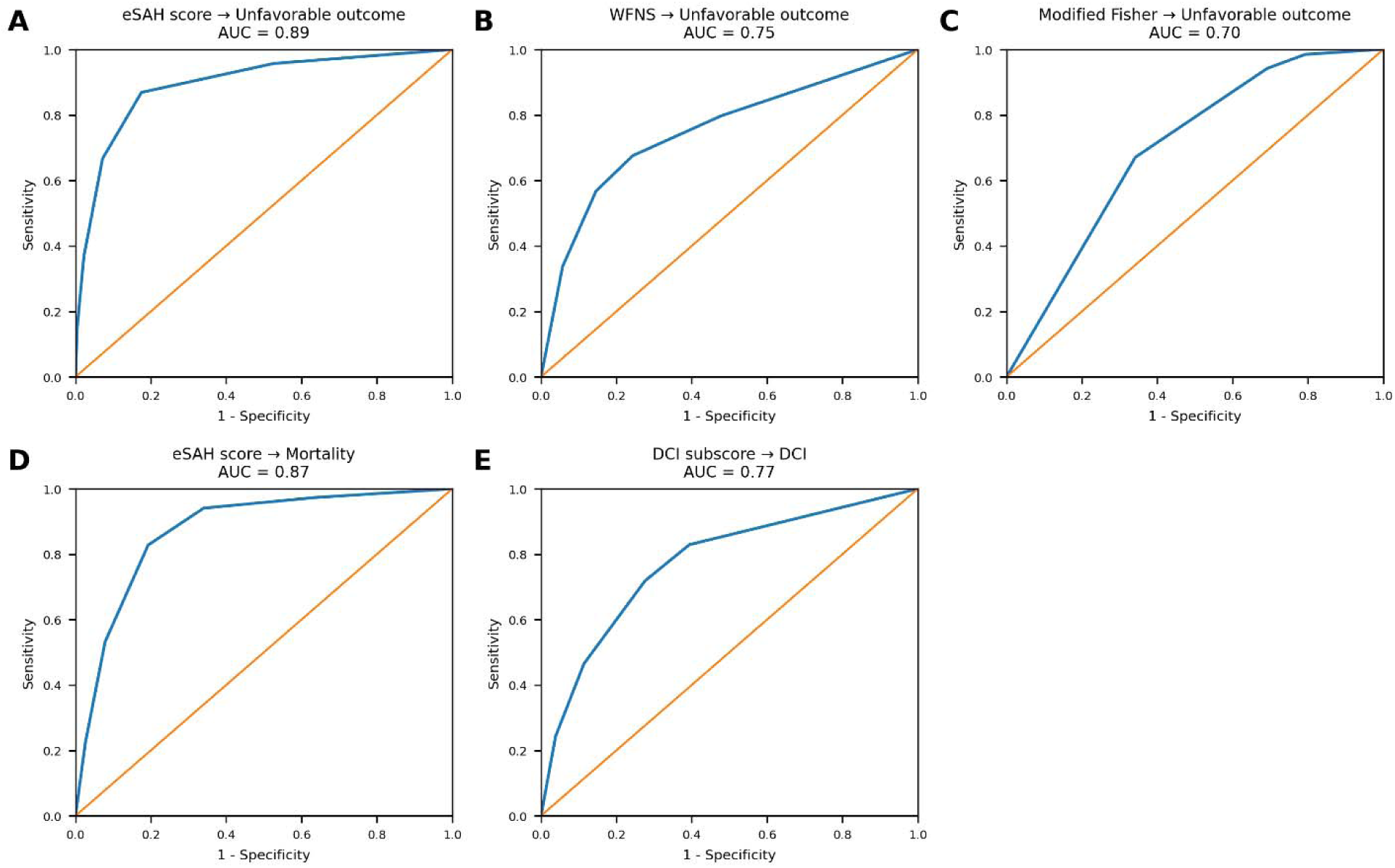
ROC curves compared to outcome measures in patients with aSAH. (A) ROC curve for eSAH score predicting unfavorable outcome (AUC = 0.89). (B) ROC curve for WFNS score predicting unfavorable outcome (AUC = 0.75). (C) ROC curve for modified Fisher score predicting unfavorable outcome (AUC = 0.70). (D) ROC curve for in-hospital mortality based on eSAH score (AUC = 0.87). (E) ROC curve for DCI based on DCI subscore (AUC = 0.77). aSAH indicates aneurysmal subarachnoid hemorrhage; AUC, area under the curve; DCI, delayed cerebral ischemia; eSAH, enhanced subarachnoid hemorrhage; ROC, receiver operating characteristic; WFNS, World Federation of Neurosurgical Societies.

Outcome distributions across eSAH score categories are illustrated in Figure 2. Increasing eSAH scores were associated with a progressive increase in both mortality and unfavorable functional outcomes.

**Figure 2:**
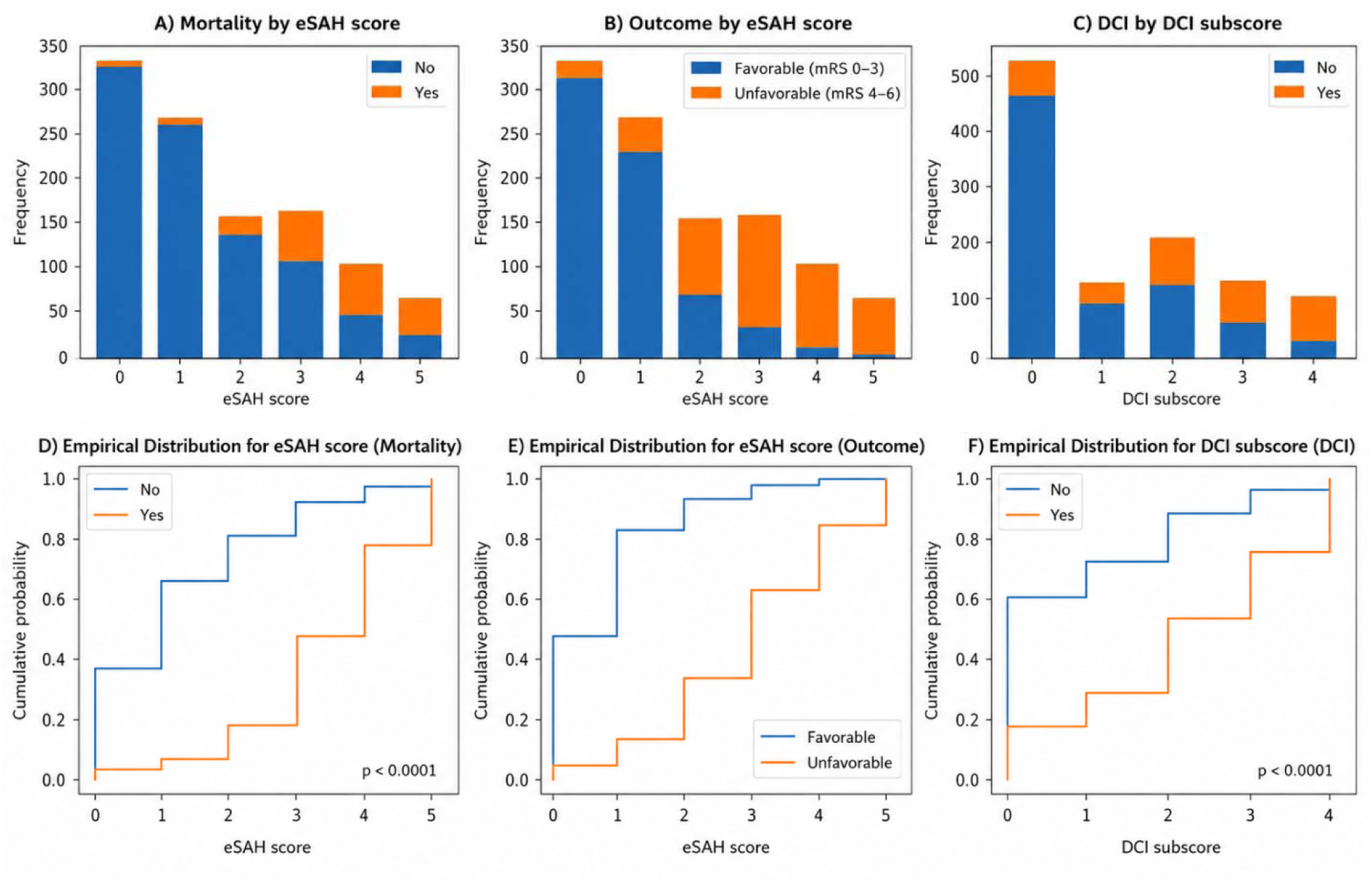
**Distribution of Outcomes Across eSAH Score and DCI Subscore Categories.**

As shown in Figure 3A, mortality increased substantially with rising eSAH scores, with very low mortality among patients with scores of 0–1 and markedly higher mortality in patients with scores ≥4. Similarly, the proportion of unfavorable functional outcomes increased steadily with increasing eSAH score (Figure 3B), indicating a strong relationship between score severity and clinical outcome.

The distribution of DCI according to DCI subscore categories is shown in Figure 3C, demonstrating higher frequencies of DCI with increasing subscore values.

Empirical cumulative distribution plots further illustrate the separation of score distributions between outcome groups (Figure 3D-F). Patients with mortality or unfavorable outcomes exhibited significantly higher eSAH scores compared with survivors or patients with favorable outcomes (p<0.0001). A similar shift toward higher scores was observed in patients who developed DCI compared with those who did not.

Detailed outcome distributions according to eSAH score are shown in Table 2. Increasing eSAH scores were associated with progressively higher rates of unfavorable outcome and mortality. Favorable outcomes were predominant among patients with scores 0-1, whereas higher scores were associated with markedly worse outcomes, with mortality exceeding 60% in patients with scores ≥4.

**Table 2:** Distribution of Functional Outcome and Mortality Across eSAH Score Categories.

| **eSAH_score** | **No.** | **Favorable,**<br>**No. (%)** | **Unfavorable,**<br>**No. (%)** | **Mortality, No,**<br>**No. (%)** | **Mortality, Yes,**<br>**No. (%)** |
| --- | --- | --- | --- | --- | --- |
| 0 | 335 | 317 (94.6) | 18 (5.4) | 330 (98.5) | 5 (1.5) |
| 1 | 271 | 234 (86.3) | 37 (13.7) | 265 (97.8) | 6 (2.2) |
| 2 | 154 | 69 (44.8) | 85 (55.2) | 133 (86.4) | 21 (13.6) |
| 3 | 158 | 33 (20.9) | 125 (79.1) | 103 (65.2) | 55 (34.8) |
| 4 | 104 | 12 (11.5) | 92 (88.5) | 47 (45.2) | 57 (54.8) |
| 5 | 66 | 3 (4.5) | 63 (95.5) | 24 (36.4) | 42 (63.6) |

The distribution of DCI according to DCI subscore is presented in Table 3. Higher DCI subscores were associated with increased frequency of DCI, indicating moderate risk stratification across subscore categories.

**Table 3:** Distribution of DCI According to DCI Subscore Categories.

| **DCI_subscore** | **No.** | **DCI, No,**<br>**No. (%)** | **DCI, Yes,**<br>**No. (%)** |
| --- | --- | --- | --- |
| 0 | 552 | 468 (89.7) | 54 (10.3) |
| 1 | 126 | 91 (72.2) | 35 (27.8) |
| 2 | 205 | 125 (61.0) | 80 (39.0) |
| 3 | 128 | 58 (45.3) | 70 (54.7) |
| 4 | 107 | 30 (28.0) | 77 (72.0) |

## Discussion

The findings in our multicenter cohort confirm the robust external validity of the original model and support its preserved reproducibility when applied outside the derivation setting^15^. In this study of 1,088 patients across three tertiary care centers, the eSAH score demonstrated excellent discrimination for unfavorable functional outcome and in-hospital mortality. While the DCI subscore demonstrated good discriminatory performance for delayed cerebral ischemia. Notably, discrimination in this validation cohort was not attenuated compared with the original study^15^, supporting the reproducibility and stability of the score across tertiary care environments.

A major finding is the comparative performance of the eSAH score relative to the established grading systems such as the WFNS and the mFs. Our eSAH score demonstrated superior predictive performance for unfavorable functional outcome. While WFNS and mFS remain used in practice, both systems rely on semiquantitative and examination based assessments that are affected by factors such as observer variability, sedation, hydrocephalus, metabolic abnormalities and systemic illness. In contrast, the eSAH score integrates neurological examination with quantitative hemorrhage volume providing a more objective and reproducible assessment of disease severity.

Early risk stratification in aSAH remains critical at the bedside. Current guidelines emphasize rapid aneurysm securing, nimodipine administration, and close neurocritical care monitoring^1,4^. However, other decisions regarding triage, interhospital transfer, intensive monitoring, and escalation of care often occur in the emergency department and may extend into the early ICU phase, especially when prognostic uncertainty is high. Hence, the strong discrimination of the eSAH score can function as an early bedside severity classifier using admission variables. Given evidence that treatment at high-volume centers and within trained NICU teams is associated with improved outcomes^17,18^, an objective and reproducible severity score may support transfer decisions and resource allocation across stroke systems of care. Further structured prognostic frameworks can help mitigate bias during the early ICU phase of care^19,20^.

The strong performance of the eSAH score reflects the integration of complementary domains of disease severity. Admission GCS remains a strong predictor of outcome in aSAH and has formed the basis of grading systems^5^. Radiographic hemorrhage burden has also been associated with vasospasm risk and functional outcomes^21–23^. Quantitative measurement of SAHV using an ABC/2-derived approach provides continuous volumetric resolution beyond conventional scales^24,25^. Hence, this validation supports hemorrhage volume as a marker of severity.

Compared with existing prediction models, the observed discrimination compares favorably with prior tools. Parekh et al. systematically reviewed heterogeneity across aSAH prediction models including variability in predictor selection and limited external validation^26^. Many models incorporate complex variables that limit bedside feasibility. In contrast, the eSAH score relies on only three admission parameters, supporting its practicality.

Notably, the DCI subscore demonstrated good discriminatory performance with an AUC of 0.77, but this is consistent with the multifactorial nature of DCI^15^. However, while machine learning approaches that integrate larger datasets have shown improved DCI prediction, these models often require more complex variables.

The outcome distribution analyses further support the clinical validity of the model. Increasing eSAH scores were associated with progressively higher rates of mortality and unfavorable functional outcomes, while higher DCI subscores correspond to increasing frequencies of delayed cerebral ischemia. The cumulative distribution plots demonstrated clear separation between favorable and unfavorable outcome groups, supporting the discriminatory capacity of the scoring system across clinical endpoints.

Quantitative severity stratification has implications for economic planning and healthcare resource allocation, as aSAH is associated with substantial hospitalization burden ^8,19,27^. Advances in automated segmentation and AI based volumetric technology may facilitate implementation of quantitative approaches such as the eSAH score ^13^. Recent evidence suggests that structured collaboration between clinicians and artificial intelligence (AI) enhances reproducibility and clinical integration^7,28,29^.

Our study has several limitations. Although the cohort was substantially larger and multicenter the study remained retrospective in design. In addition, long term functional outcomes beyond hospital discharge, such as mRS outcomes after 90 days were not included, as the primary purpose of the eSAH score was early prediction of discharge outcomes and in-hospital mortality. Future prospective studies with longitudinal follow up to evaluate long term functional outcomes. Nevertheless, strict adherence to the original soring metrics strengthens the validity of this independent external evaluation. Collectively, these findings support the integration of age, neurological examination, and quantitative hemorrhage burden for early stratification of aSAH severity.

## Conclusion

The eSAH score demonstrates strong external validity and reproducibility in an independent cohort. Further studies are needed to support its integration with AI-based volumetric imaging analyses.

## Data availability statement

Deidentified patient data that support the findings of this study are not publicly available due to privacy and institutional restrictions but can be made available from the corresponding author upon reasonable request. All other relevant data are included in the manuscript.

